# Factors influencing SARS-CoV-2 IgG test sensitivity: A Bayesian analysis of seroconversion and reversion by time since infection, test, age and disease severity

**DOI:** 10.1101/2025.06.29.25330529

**Authors:** Toon Braeye, Steven Abrams, Niel Hens

## Abstract

1.

**BACKGROUND:** Antibody testing is commonly used to assess past exposure to pathogens, but the interpretation is complex. We quantified test-specific SARS-CoV-2 seroconversion and reversion by time since PCR-confirmed infection, age and disease severity.

**METHODS:** We used Belgian data from laboratory SARS-CoV-2 testing, prescriptions, contact tracing and hospital surveillance collected between March 2020 and June 2021. Additionally, we gathered data for the Wantai and EuroImmun IgG serological tests from the scientific literature.

We used a hierarchical Bayesian model to estimate distributional parameters of a scaled Weibull-bi-exponential distribution for the time-varying sensitivity of qualitative serological test results obtained after PCR-confirmed SARS-CoV-2 infection. We accounted for disease severity (distinguishing between asymptomatic, symptomatic, and hospitalized cases), age (i.e., in terms of age groups 18-49, 50-64, and 65-74 years) and serological test used.

**RESULTS:** We included 44,262 serological test results: 10,864 obtained from published studies, 33,398 from Belgian laboratories. Seroconversion occurred during the six weeks following a PCR-confirmed infection. For the EuroImmun test, 82% (95%CrI: 80%-84%) of symptomatic individuals in the youngest age group seroconverted, compared to 95% (95%CrI: 95%-96%) for the Wantai test. In addition, seroconversion was associated with hospitalization (OR = 6.98, 95%CrI: 4.85-11.37, compared to asymptomatic infection) and older age (OR = 1.67, 95%CrI:1.43-1.92, for 65-74-year-olds compared to 18-49-year-olds). Reversion after initial seroconversion was strongly associated with the test used. At 50 weeks, the proportion of symptomatic individuals aged 18-49 years who remained seropositive was 63% (95%CrI: 56%-69%) for the EuroImmun test and 95% (95%CrI: 94%-96%) for the Wantai test. Slower reversion was associated with severe infection and older age.

**CONCLUSION:** Seropositivity after SARS-CoV-2 infection was significantly associated with the type of test used, age of the case and severity of the infection. More severe infection and older age resulted in increased and prolonged seropositivity.

## 2. Introduction

Starting in 2020, numerous seroprevalence studies were conducted to understand the extent of past exposure to SARS-CoV-2 within different populations [1]. These studies used immunoassays, serological tests, capable of directly or indirectly detecting antibodies. These antibodies typically target one of the four structural proteins of the SARS-CoV-2 virus: spike (S), membrane (M), envelope (E), or nucleocapsid (N) protein. The S protein, which is further divided into the N-terminal domain (NTD) and the receptor-binding domain (RBD), along with the N protein, are the primary immunogens and are typically the targets of immunoassays [2]. Assays determine the amount of antibodies and manufacturer suggested threshold values can be used to translate quantitative to qualitative results. Qualitative results are then typically reported as the proportion of positive samples among all samples. The interpretation of such results however is not straightforward as immunoassays are imperfect. Qualitative results are associated with a proportion of false negatives (sensitivity below 100%) and false positives (specificity below 100%) [3]. In addition, the main research interest is often not solely in the presence of antibodies. The main objective of seroprevalence studies, in addition to objectives regarding susceptibility, concerns the proportion of persons previously infected: the cumulative infection rate. The sensitivity with respect to previous infection, as compared to sensitivity to a certain level of antibodies, is a dynamic metric which will depend on the test, but also on antibody kinetics associated with the time since infection and person and disease characteristics [3].

Quantifying this dynamic sensitivity is difficult as it is affected by different factors. We briefly introduce the three main factors: (A) how infections are diagnosed, (B) which persons are included in the cohort under follow-up and (C) how is time since infection included in the analysis. With respect to (A): studies typically use either a ‘gold standard’ immunoassay or a combination of immunoassays, neutralization assays or RT-PCR tests to determine ‘true’ positivity of an individual [4]. The shortcoming of any of these options typically boils down to the gold standard’s own sensitivity and specificity. (B) Patients included in longitudinal studies were frequently hospitalized. Hospitalization is typically associated with high antibody titers not reflective of titers in asymptomatic patients [5–7]. Metareviews reported a high risk of patient selection bias in 97-98% of assessments [4,8]. (C) After an initial increase during 3 to 12 weeks for SARS-CoV-2 [8], antibody titers will decrease again over time, while antibody avidity and affinity might increase. The decline is not monophasic. An initial strong decrease is followed by a plateau [9,10]. Whether or not this decrease is antigen-specific is still under debate. Antibodies associated with S1 seem more durable than those associated with the N-protein [2,11–16]. Furthermore, the decrease is linked to patient selection. The half-time of IgG S-protein titers associated with asymptomatic cases is less than half of that of mild cases and has been estimated at 55 days [17,18] or even shorter (i.e., 36 days as estimated by Ibarrondo et al. [19]). No detectable neutralizing activity was found in 50% of asymptomatic infections 1 year after infection [10]. The role of sex and age is less clear [20]. More stable antibody levels have been reported for females [21]. Studies have reported higher initial titer concentrations in older age groups. Higher initial titers have been linked to extended seropositivity [7,14,21–23]. Typical methods to adjust for the imperfect performance of serological tests, such as a Rogan-Gladen type estimator [24], can be extended to include time-varying sensitivity estimates, but with the exception of some modelling studies, seroprevalence studies typically do not correct for waning of antibodies [25].

We aim to estimate distributional parameters associated with an underlying parametric distribution for time-varying, test-specific sensitivity in relation to time since infection and to quantify the effect of age and clinical severity on this distribution. While estimating the sensitivity, all available data, including estimates obtained from the literature, will be included. Such an estimate for the time-varying and test-specific sensitivity is necessary for a meaningful interpretation of qualitative individual serological test results and for the translation of population-level seroprevalence results to (cumulative) incidence estimates.

## 3. Methods

We used a Bayesian hierarchical model with a binomial likelihood to fit the number of positive serological tests out of all serological tests at week 𝑡 after PCR-confirmation of SARS-CoV-2 infection. Data on cases was collected from Belgian laboratories and published studies. We first present the distributions for seroconversion and reversion and how these processes result in a temporal state of detectable antibodies, coined seropositivity.

### 3.1. MODEL STRUCTURE

We modeled two immunological processes: seroconversion and seroreversion. Seroconversion is the process of reaching a detectable level of antibodies after infection. Seroreversion is the process of losing that detectable level since having first attained it. These processes are represented by two random variables: 𝑇_𝑐_denotes the time between infection and conversion and 𝑇_𝑟_is the time between conversion and reversion. 𝑇_𝑐_is assumed to follow a Weibull distribution. 𝑇_𝑟_ follows a bi-exponential distribution. The Weibull distribution was selected because it can flexibly model time-to-event data. The bi-exponential distribution allowed us to include a mixture of a fast and slow decrease. Four parameters representing different time-to-event processes need to be estimated: a scale and shape parameter for 𝑇_𝑐_and two exponential parameters for 𝑇_𝑟_. In addition, we need to estimate the proportion of cases that undergo seroconversion and a parameter that determines the mixture between slow and fast decrease.

The random variable 𝑆_𝑡_ (with discrete probability density function ( h(𝑆_𝑡_)) represents the proportion of seropositive cases at week 𝑡 out of all cases infected at week 1. To account for both the time to seroconversion, the time to seroreversion given seroconversion and the overall proportion that will seroconvert we include h(𝑆_𝑡_) as (notation based on Shioda et al. [26]):

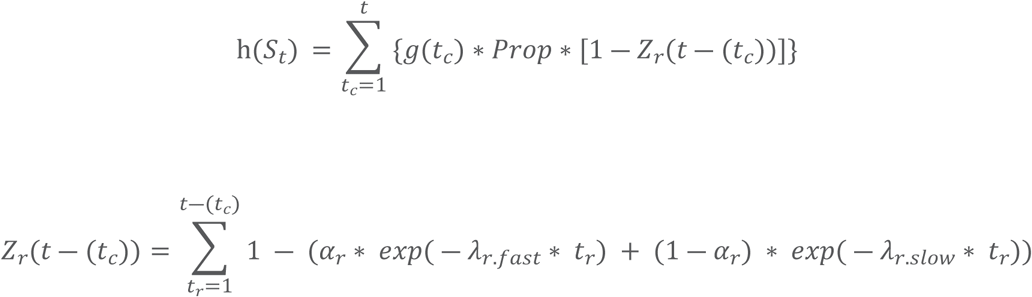

For a case to be seropositive at week 𝑡 after infection, it had to undergo seroconversion at week 𝑡_𝑐_ with 𝑡_𝑐_ before or at week 𝑡. 𝑔(𝑡_𝑐_) is the discretized probability density function for 𝑇_𝑐_. In addition, the case should not have undergone seroreversion during the time interval 𝑡 ― (𝑡_𝑐_). 𝑍_𝑟_ denotes the cumulative density function of 𝑇_𝑟_. 𝑍_𝑟_(𝑡 ―(𝑡_𝑐_)) is the proportion of cases seroconverted at week 𝑡_𝑐_ that will have undergone seroreversion by or at week 𝑡. Seroreversion will only occur given seroconversion, we therefore have to scale 𝑍_𝑟_(𝑡 ―(𝑡_𝑐_)): 𝑔(𝑡_𝑐_) ∗ 𝑃𝑟𝑜𝑝 ∗ 𝑍_𝑟_(𝑡 ―(𝑡_𝑐_)).

The parameters for the time-to-event distributions (𝑠𝑐𝑎𝑙𝑒_𝑐_, 𝑠ℎ𝑎𝑝𝑒_𝑐_, 𝜆_𝑟.𝑓𝑎𝑠𝑡_, 𝜆_𝑟.𝑠𝑙𝑜𝑤_) are global. The proportion, 𝑃𝑟𝑜𝑝, converted and the seroreversion-mixture, 𝛼_𝑟_, are group-specific with the following linear predictors.

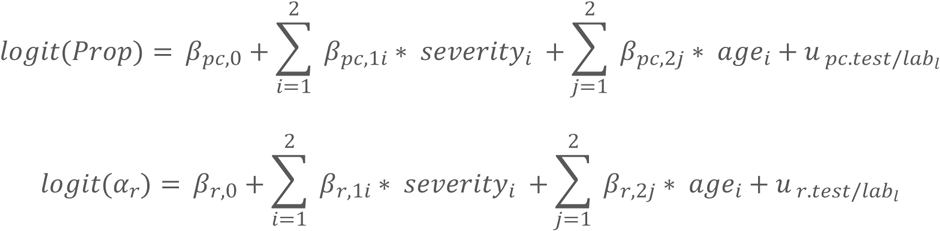

There are three age groups and three severity groups. Coefficients for the first groups are set to zero, leaving two coefficients to be estimated per distributional parameter. 𝑢 _𝑡𝑒𝑠𝑡/𝑙𝑎𝑏𝑙_ represent the random effect for tests and laboratories. We evaluated multiple approaches for modelling antibody reversion kinetics, including linear decay functions and various time-to-event distributions such as Weibull. After comparative analysis, we selected a bi-exponential distribution. This model optimally captured the biphasic nature of antibody dynamics: an initial rapid decline followed by a slower, plateau-like phase. While 𝛼_𝑟_ was allowed to vary by severity, age group, and test, we determined through model selection that the decay rate parameters 𝜆_𝑟.𝑓𝑎𝑠𝑡_ and 𝜆_𝑟.𝑠𝑙𝑜𝑤_ could be treated as global parameters without significant loss of fit. Multiple model structures were evaluated using Bayesian Information Criterion (BIC) and convergence diagnostics, including interaction terms.

We used non-informative priors for the model coefficients (i.e., normal distributions with a relatively large standard deviation of 100). The factors for test (or lab) were included as random effects with mean zero and a gamma distributed prior for the standard deviation. Markov Chain Monte Carlo (MCMC) sampling was performed using the R package *nimble*. We used three MCMC chains with 10,000 iterations each and a burn-in of 4,000 iterations to perform posterior inference, while convergence of the different chains was checked using the Gelman-Rubin statistic.

### 3.3. LABORATORY DATA

Reporting of laboratory results to a centralized database was mandatory during the COVID-19 pandemic in Belgium. While SARS-CoV-2 IgG testing was not required in any situation, a large number of IgG tests was performed. Within the centralized databases available through the LinkVacc project, test results could be linked with data on vaccination, contact tracing and hospital surveillance using pseudologized unique identifiers. We grouped test results by severity, age, test/laboratory and number of weeks since PCR-confirmed infection. The number of positive samples out of all samples is fitted to the previously defined binomial likelihood.

We considered three age groups: 18 to 49 years, 50 to 64 years and 65 to 74 years and three different categories of clinical severity: self-reported asymptomatic, self-reported symptomatic or notified hospitalized. Symptoms could be reported either before diagnosis (e.g., during the consultation) or during contact tracing (when cases were interviewed about their contacts). If the case reported symptoms at any point, the case was classified as symptomatic. If, upon investigation, the case reported no symptoms, it was classified as asymptomatic. If this information was missing, the records were excluded. Additionally, if a hospitalization for COVID-19 was reported in a period of 7 days before to 60 days after the first positive PCR test through the clinical hospital survey, the case was classified as hospitalized [27]. Unfortunately, the specific serological test and cut-offs used were not reported by the laboratory. The reporting laboratory was therefore used as a substitute for test and included as a random variable in the model formulated in Section 3.1.

We only included information on subjects with positive PCR tests conducted in 2020, given the documented changes in epidemiology and immunological responses after infection with new variants (as compared to the wild-type SARS-CoV-2 strain) emerging from 2021 onwards. We excluded all IgG tests that followed any subsequent PCR test after the first positive test as PCR-testing could be indicative for additional exposure to SARS-CoV-2. IgG test results after vaccination were also excluded from the analysis. We included no IgG test results obtained after June 2021.

### 2.3. LITERATURE DATA

We included two tests, chosen because of their use in two repeated cross-sectional seroprevalence studies in Belgium, a semi-quantitative test (EuroImmun), targeting the S1 protein and a semi-quantitative test (Wantai) targeting the RBD. We included all studies listed in the systematic review by Owusu-Boaitey et al. [3] in our statistical analysis. For the Wantai test, we also included the findings from the study by Hønge et al. [28], originally excluded by Owusu-Boaitey et al. because the study cohort, consisting exclusively of healthy blood donors, was not considered representative. Since our model allowed us to differentiate by disease severity (including asymptomatic presentation), we did include the study.

The data from scientific literature is included in the model in a similar way to the data obtained from Belgian laboratories. As opposed to the data from the Belgian laboratories, the specific test is known and included in the model as level for the random effect 𝑢 _𝑡𝑒𝑠𝑡/𝑙𝑎𝑏𝑙_. For each included paper, we obtained the test, the number of (positive) samples by weeks since infection, the proportions of asymptomatic, symptomatic and hospitalized persons in the cohort and the proportions within the age groups 18-49, 50-64 and 65-74 years. Whenever specifics on severity and age were not available, we included the following default distributions. For clinical severity: 50% asymptomatic, 45% symptomatic and 5% hospitalized (severe) as this was reported by Takahashi et al. [29] to be the average over serosurveys. For age: we either included the default for Germany, if the study was performed in Germany: 18-49 (45%), 50-64 (35%) and 65-74 (20%) based on Neuhauser et al. [30] or the default for Denmark: 18-49 (64%), 50-64 (26%) and 65-74 (10%) based on Pires et al. [31]. For studies and regions outside of Denmark and Germany for which we could not obtain a specific age distribution, we used as default a distribution in between the German and Danish distributions: 18-49 (55%), 50-64 (30%) and 65-74 (15%). Details by study and weeks since infection are provided in S1 Table.

## 4. Results

### 4.1. NUMBERS INCLUDED

In the centralized database there were 472 223 persons with a positive PCR test in 2020 in Belgium in the age group from 18 years to 74 years. Of these persons 15% (N = 70 951) had IgG tests (N = 93 127) before July 2021. We had to exclude (in order of exclusion criterion) 6 856 IgG tests because they followed vaccination, 39 622 tests because they either preceded the first positive PCR test or followed any PCR test after the first positive PCR test. For 13 251 IgG tests, data on the severity of infection was missing. Therefore, we could include 33 398 IgG tests from 30 002 persons reported by 84 laboratories. The laboratory with the most records accounted for 14% of all IgG tests.

Of these tests 28% were taken from individuals with asymptomatic infection, 67% from individuals with symptomatic infection and 5% from hospitalized individuals. The division by age group was 51% (18-49 years), 35% (50-64 years) and 14% (65-74 years). 16% of tests were collected in the first 4 weeks, 42% in weeks 4-11, 40% in weeks 12-35 and 1% later. The numbers by weeks since first positive PCR test, clinical severity and age group are presented in Table 1.

**Table 1:** Number (and percentage) of included IgG tests following a positive SARS-CoV-2 PCR test in 2020, by weeks since positive PCR test, clinical severity and age group, Belgian laboratory data.

| Severity | Weeks since positive PCR test | 18-49 years | 50-64 years | 65-74 years |
| --- | --- | --- | --- | --- |
| Asymptomatic | 0-3 | 1247 (3.7) | 687 (2.1) | 285 (0.9) |
|  | 4-11 | 1854 (5.6) | 1139 (3.4) | 506 (1.5) |
|  | 12-35 | 1917 (5.7) | 1083 (3.2) | 400 (1.2) |
|  | 36+ | 145 (0.4) | 50 (0.1) | 8 (0) |
| Symptomatic | 0-3 | 1430 (4.3) | 965 (2.9) | 421 (1.3) |
|  | 4-11 | 5125 (15.3) | 3520 (10.5) | 1430 (4.3) |
|  | 12-35 | 5063 (15.2) | 3357 (10.1) | 1082 (3.2) |
|  | 36+ | 64 (0.2) | 33 (0.1) | 4 (0) |
| Hospitalized | 0-3 | 102 (0.3) | 135 (0.4) | 162 (0.5) |
|  | 4-11 | 129 (0.4) | 288 (0.9) | 198 (0.6) |
|  | 12-35 | 121 (0.4) | 266 (0.8) | 167 (0.5) |
|  | 36+ | 5 (0) | 4 (0) | 6 (0) |

### 4.2. HIERARCHICAL BAYESIAN MODEL

#### 4.2.1. Group specific effects

The proportion that eventually undergoes seroconversion was associated with test, disease severity and age. More specifically, the odds ratio of seroconversion when individuals were eventually hospitalized as compared to self-reported asymptomatic was 6.98 (95%CrI: 4.85-11.37). Higher seroconversion was associated with older age (OR 1.67 95%CrI 1.43-1.92) compared to the youngest age group (18-49-years-old) (Table 2). Faster waning was associated with the youngest age group and symptomatic infection as compared to the older age groups and asymptomatic infection (Table 3). The effect of the different tests/laboratories is presented in the supporting information.

**Table 2:** Odds Ratios (OR) derived from the posterior distributions of the regression coefficients for the proportion that eventually undergoes seroconversion after a positive PCR-test in 2020 (asymptomatic = self-reported asymptomatic, symptomatic = self-reported symptomatic, hospitalized = notified hospitalized, SD = standard deviation), Belgian laboratory data and data from published research.

| Variable | OR | 95% CrI: |  |
| --- | --- | --- | --- |
|  |  | Lower | Upper |
| Age |  |  |  |
| 18-49 (ref) | 1 |  |  |
| 50-64 | 1.31 | 1.16 | 1.47 |
| 65-74 | 1.67 | 1.43 | 1.92 |
| Severity |  |  |  |
| Asymptomatic (ref) | 1 |  |  |
| Symptomatic | 2.21 | 1.95 | 2.55 |
|  |  | Lower | Upper |
| Hospitalized | 6.98 | 4.85 | 11.37 |
| Random intercept | SD |  |  |
| test/lab | 1.71 | 1.51 | 1.97 |

**Table 3:** Odds Ratios (OR) derived from the posterior distributions of the regression coefficients for the proportion associated with faster waning (asymptomatic = self-reported asymptomatic, symptomatic = self-reported symptomatic, hospitalized = notified hospitalized, SD = standard deviation), Belgian laboratory data and data from published research.

| Variable | OR | 95% CrI: |  |
| --- | --- | --- | --- |
|  |  | Lower | Upper |
| Age |  |  |  |
| 18-49 (ref) | 1 |  |  |
| 50-64 | 0.12 | 0.02 | 0.29 |
| 65-74 | 0.03 | 0 | 0.14 |
| Severity |  |  |  |
| Asymptomatic (ref) | 1 |  |  |
| Symptomatic | 2.58 | 1.23 | 6.91 |
| Hospitalized | 0 | 0 | 0.62 |
|  |  | Lower | Upper |
| Random intercept | SD |  |  |
| test/lab | 2.41 | 1.58 | 3.69 |

#### 4.2.2. SeroConversion and SeroReversion

We present the posterior distributions for seroconversion and seroreversion separately by clinical severity and age group for the EuroImmun and Wantai test in Fig 1. The process of seroconversion was set equal over groups. By week 5, over 95% of those that will seroconvert had seroconverted. The seroreversion occurred faster in younger age groups and in those with less severe infections not requiring hospitalization. The EuroImmun test was associated with more and faster seroreversion compared to the Wantai test. The combined effects of proportion, conversion and reversion over time since PCR-confirmed infection are presented in Fig 2.

**Fig 1:**
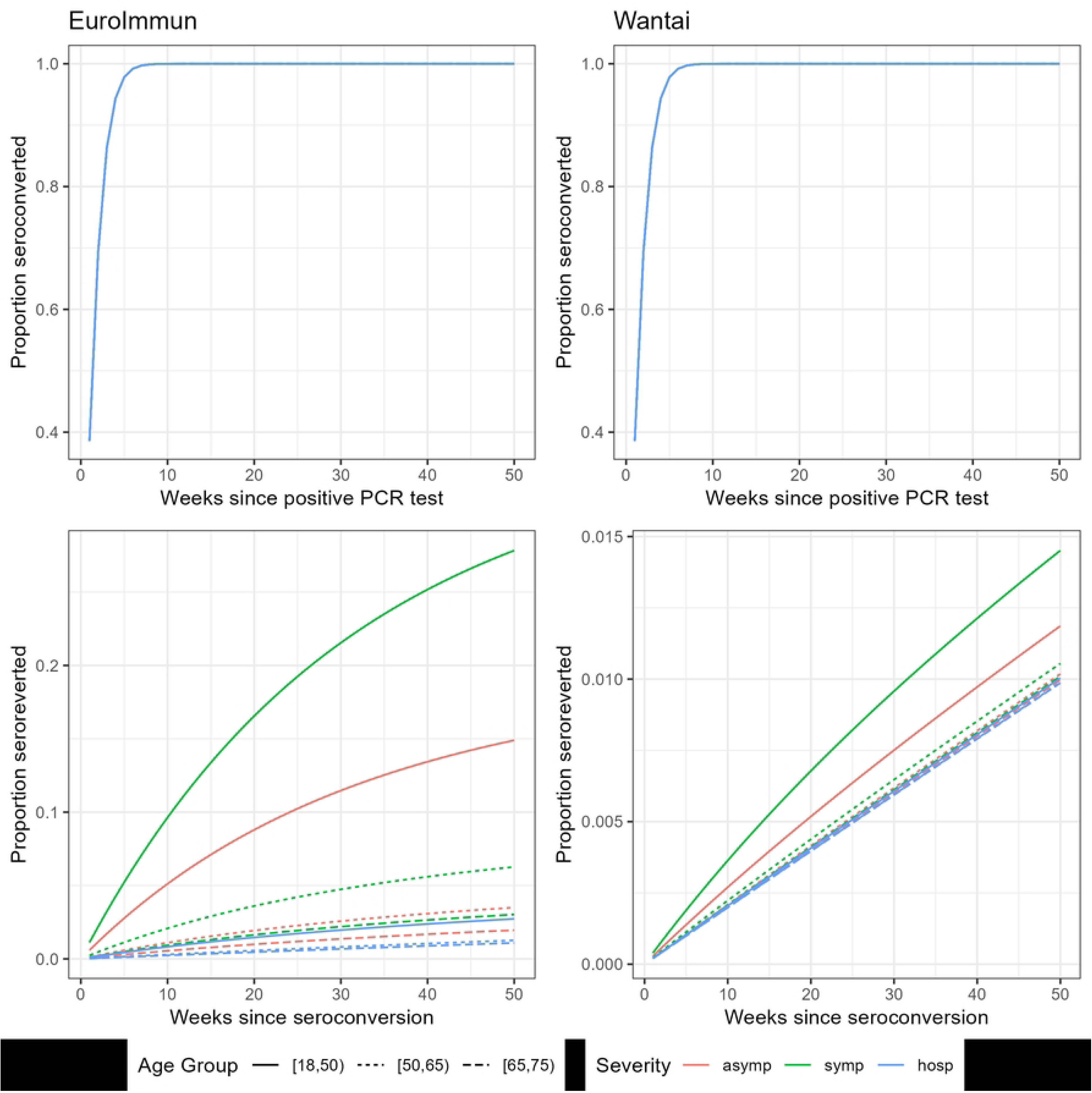
Plots of the posterior Weibull distributions for seroconversion (upper) and seroreversion (lower) by weeks since positive PCR test, clinical severity and age group for the EuroImmun (left) and the Wantai (right) test, Belgian laboratory data and data from published research.

**Fig 2:**
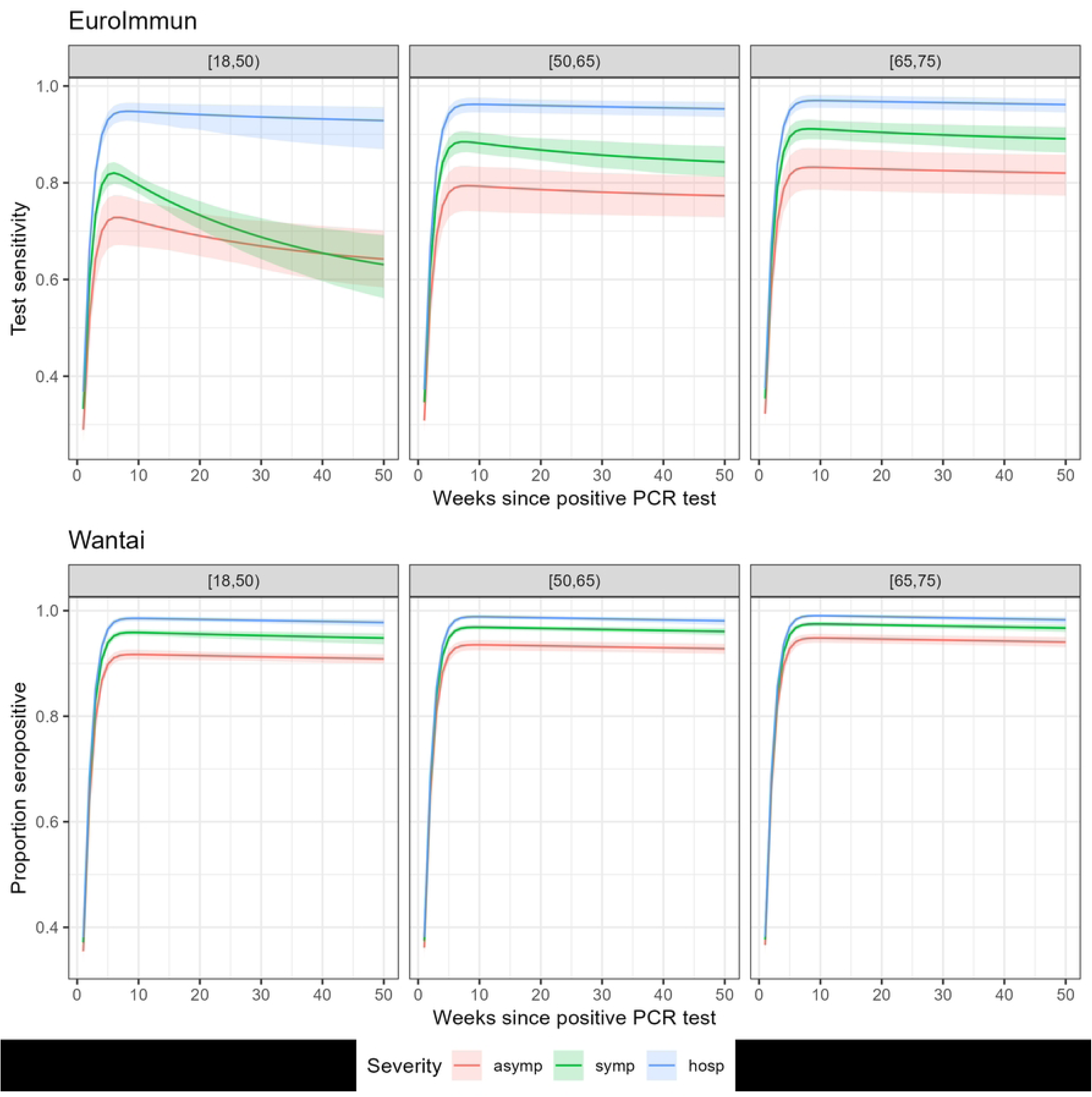
Plots of the posterior scaled Weibull-Bi-exponential distribution for time-varying seropositivity for the EuroImmun (upper) and Wantai (lower) test by weeks since positive PCR test, clinical severity and age group, Belgian laboratory data and data from published research.

#### 4.2.3. Goodness of fit

We present the model fit to both Belgium’s laboratory data (Fig 3) and the data obtained from literature (Fig 4). Considerable variation was linked to the test used or reporting laboratory. We presented the distributions of the random effects (scale reversion and proportion converted) in S1 Fig.

**Fig 3:**
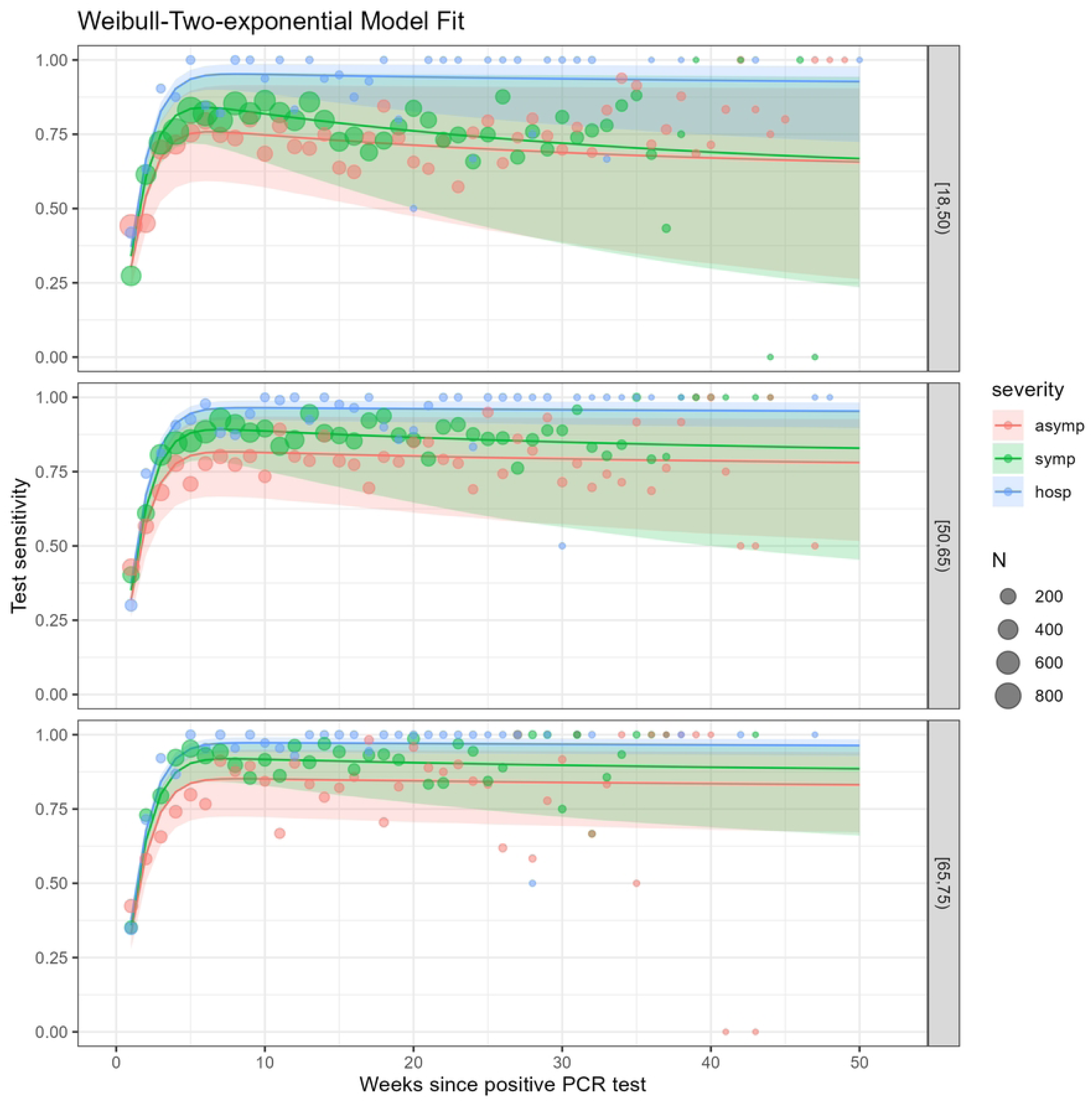
Plots of the posterior scaled Weibull-Wi-exponential distribution for the time-varying seropositivity averaged over all laboratories (unweighted average) by weeks since positive PCR test, clinical severity and age group, Belgian laboratory data, IgG tests after a positive PCR test in 2020.

**Fig 4:**
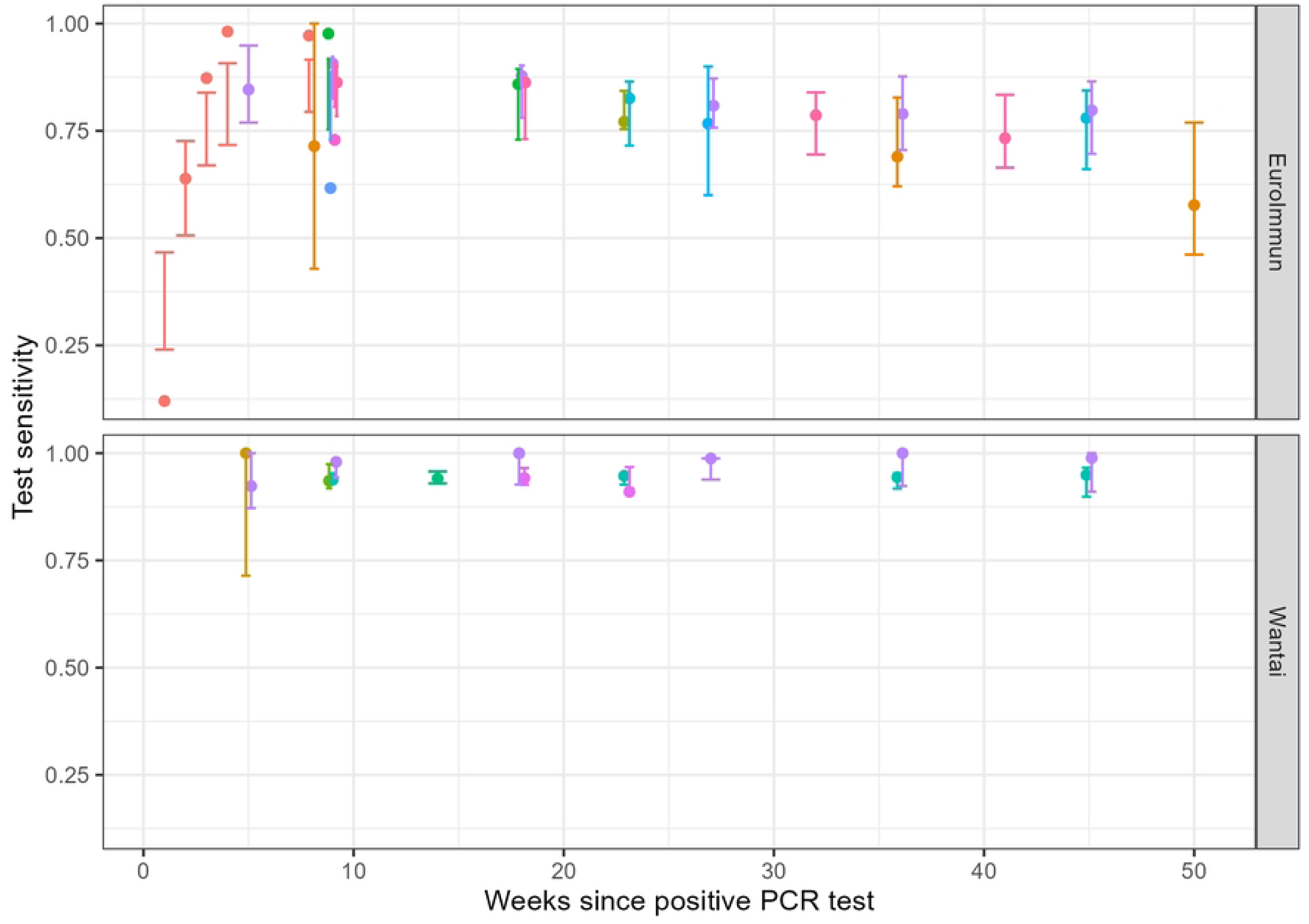
Posterior Predictive Checks: The proportion positive reported (dot) and the posterior binomial confidence interval (error bars) by study (color) and weeks since positive PCR test for the EuroImmun (upper) and Wantai (lower) test, data from published research.

## 5. Discussion

This study provides a comprehensive analysis of factors influencing SARS-CoV-2 IgG test sensitivity using a hierarchical Bayesian approach including data from published studies and Belgian laboratories. Our key findings demonstrate that seropositivity following SARS-CoV-2 infection is significantly influenced by three primary factors: the specific serological test used, the age of the individual, and the severity of the initial infection.

Adjusting qualitative seroprevalence results for test-specific sensitivity and specificity to estimate past exposure has become more common, but remains relatively rare. In a 2020 systematic review of global SARS-CoV-2 antibody seroprevalence, Bobrovitz et al. reported that only 24% of studies provided sensitivity and specificity estimates, with an even smaller proportion adjusting their seroprevalence estimates accordingly [32]. The importance of these adjustments has been demonstrated. Studies using the same seroprevalence data, but different sensitivity estimates have reported considerably different infection fatality rates [33,34]. The limited use may be attributed to the complexity of these metrics. Sensitivity depend not only on the test itself but also on the study cohort and time since infection. For example, for the EuroImmun serological test sensitivity estimates have been reported ranging from 94% to 53%: the manufacturer reported a sensitivity ≥10 days post symptom onset of 94.4%. Researchers in South Africa estimated sensitivity at 64.1% [6], Public Health England at 72%, 74.5% [35], 77.2% [36] and a study on Antarctic cruise passengers at 81% declining to 76% at 3 months and 53% at 1 year [37]. In this work, we propose a framework to harmonize these estimates and quantify test-specific sensitivity by time since infection and given cohort characteristics: age and severity of the infection. Previous meta-analysis with a comparable aim have disregarded these cohort characteristics and limited their included studies to those with cohorts considered representative for the general population. Given the high risk of patient selection bias associated with these studies [4,8], this is a considerable limitation. The systematic review by Owusu-Boaitey et al. [3], for example, quantified test-specific sensitivity over time but could only include one study [12] to estimate the sensitivity of the Wantai test beyond 5 months after symptom-onset. This study by Scheiblauer et al. included PCR-confirmed cases (N=390) of which none were asymptomatic and a large proportion (15.8%) were hospitalized. Other studies on the Wantai test could not be included because the cohort was not considered representative. For example, a study on blood donors [28] was excluded. We could include this population by accounting for the severity of the infection. In addition, we could include studies too recent to be included by Owusu-Boaitey et al. [28,38]. We could not include the study by Perez-Saez et al. [39] since their cohort (N=354) was established out of persons with an initial positive EuroImmun test. Not all persons within the cohort had a PCR-confirmed infection and dates of infection had to be estimated. Given initial seroconversion, they estimated 40% seroreversion after 9 months. Likewise in our study we observed a decrease in the sensitivity of the EuroImmun test over time, albeit one less rapid. We observed a drop of around 20% points for 18-49 year-olds with an initial symptomatic infection. This translates to seroreversion of around 25% of those that seroconverted over a 50 week period. For the Wantai serological test we observed both higher initial and more durable seropositivity. The assay used has previously been established as a major factor for seropositivity [40]. The Wantai serological test, the most sensitive and durable serological test in this study, remains associated with a proportion non-responders. Non-response, non-conversion or also sero-silence, is the absence of detectable antibodies at any time after infection. The proportion of sero-silence after a PCR-confirmed infection is estimated at 5.2-7.8% [41–43]. As with the previous findings this proportion also depends on the test used and the cohort characteristics.

Our other findings also largely agree with previously published longitudinal studies and reviews. The severity of infection has been associated with high and persistent antibody levels by several studies [44–49]. In addition, research reports lower sensitivity in young adults (ages younger than 40-50 years) [50–52], but the association in older age groups seems less clear with contradictory findings. Higher age has been mostly associated with slower waning [11,39,46,47,53,54] of the humoral immune response, with some studies reporting faster waning [21,48]. We found higher seropositivity in those aged over 50 as compared to those below, but the effect of age was smaller than the effect of severity of infection. We could not include persons beyond the age of 75 years. An age beyond which frailty and immunosenescence might impact sensitivity [55].

This study has several limitations. The process of seroreversion was included using a bi-exponential distribution. Some studies have opted for a Weibull distribution [26,56], others for a single exponential [57,58] or splines on the logit scale [59]. We did not extrapolate beyond the periods available within the data, but most of our data concerns the first weeks after PCR confirmation. As many other studies we set a positive PCR-test as reference. Laboratory confirmation by PCR however has its own time-varying sensitivity and specificity [60]. As no data were available on the test used by the reporting laboratory, we used the reported laboratory as level for a random effect. We opted not to explore sex because initial analysis did not associated sex with large differences in seropositivity after infection. Studies have claimed to find no strong evidence of heterogeneity in antibody persistence by sex [61]. With some claiming a more durable response in females [48]. The age groups in this analysis as well as the specific tests were chosen in preparation of future work on two cross-sectional seroprevalence studies.

In conclusion, our study provided a comprehensive framework for estimating time-varying, test-specific sensitivity in serological testing for SARS-CoV-2, accounting for the critical factors of disease severity, age, and time since infection. The results demonstrate that seropositivity is significantly influenced by test selection, with the Wantai test showing superior performance compared to the EuroImmun test in both initial detection and long-term durability. Our findings reveal that older age (50-74 year) and more severe infection lead to higher and more durable seropositivity.

## 6. Supporting Information Captions

**Fig S1:**
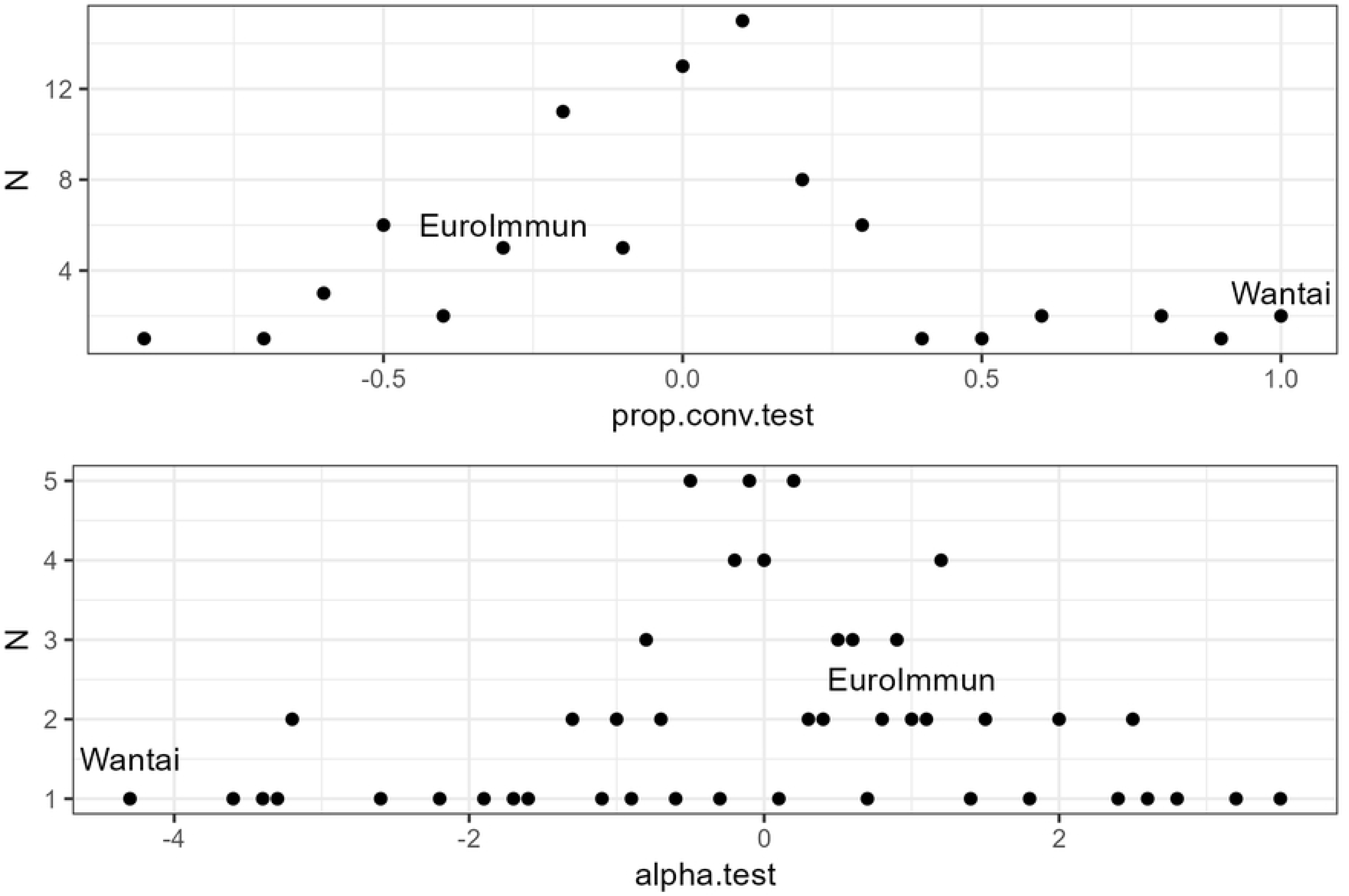
Discrete approximation of the distribution of the random effects associated with test/lab for the random effect in proportion seroconverted (upper) and the scale of the Weibull representing seroreversion (lower). The values associated with the EuroImmun and Wantai tests are annotated.

## Data Availability

Most relevant data are within the manuscript and its Supporting Information files. Additional data (COVID-19 seropositivity by time since sampling) can be requested through https://www.hda.belgium.be/en

